# Performance Characteristics of Six Immunoglobulin M (IgM) ELISA Assays Used for Laboratory Confirmation of Measles

**DOI:** 10.1101/2022.09.02.22279538

**Authors:** Sun B. Sowers, Kiana Anthony, Sara Mercader, Heather Colley, Stephen N. Crooke, Paul A. Rota, Donald R. Latner, Carole J. Hickman

## Abstract

Laboratory confirmation of infection is an essential component of measles surveillance. Detection of measles specific IgM in serum by enzyme linked immunosorbent assay (ELISA) is the most used method for confirming measles infection. ELISA formats vary as does the sensitivity and specificity of each assay. Specimens collected within 3 days of rash onset can yield a false negative result, which can delay confirmation of measles cases. Interfering substances can yield a false positive result, leading to unnecessary public health interventions. The IgM capture assay developed at the Centers for Disease Control (CDC) was compared against 5 commercially available ELISA kits for the ability to detect measles virus-specific IgM in a panel of 90 well-characterized specimens. Serum samples were tested in triplicate using each commercial kit as recommended by the manufacturer. Using the CDC measles IgM capture assay as the reference test; sensitivity and specificity for the commercial kits ranged from 50 to 83% and 86.9 to 98%, respectively. Discrepant results were observed for samples tested with all five commercial kits and ranged from 13.8 to 28.8% of the specimens tested. False positive results occurred in 2.0 to 13.1% of sera while negative results were observed in 16.7 to 50% of sera that were positive by the CDC measles IgM capture assay. Evaluation and interpretation of measles IgM serologic results can be complex, particularly in measles elimination settings. The performance characteristics of a measles IgM assay should be carefully considered when selecting an assay to achieve high quality measles surveillance.

## Introduction

Measles is a highly communicable viral infection with serious complications. Endemic measles was declared eliminated from the United States (U.S.) in 2000 due to sustained high two-dose coverage with the measles mumps rubella (MMR) vaccine (1-4). In 2019, the U.S. experienced the highest annual number of measles cases in over 25 years because of repeated introductions by U.S. residents traveling abroad and returning to under vaccinated communities within the United States. Cases were confirmed in 31 states, and outbreaks in New York City and New York State resulted in chains of transmission that continued for more than 10 months and threatened the U.S. elimination status (5, 6).

Public health and health care professionals play an important role in diagnosing and managing acute cases of measles and preventing transmission during outbreaks. Effective outbreak control relies upon rapid and accurate confirmation of measles infected individuals, interrupting chains of transmission by identification and vaccination of susceptible individuals, and by isolation and quarantine of those who are likely to spread disease following exposure. In addition, measles vaccination can provide effective postexposure prophylaxis if given within 72 h of initial exposure (7).

Laboratory confirmation of measles is an essential component of measles surveillance. Detection of measles RNA in a respiratory sample by real-time polymerase chain reaction (RT-PCR) and detection of measles-specific IgM in a serum specimen by ELISA are the two most used methods for confirming measles infection. CDC recommends collection of both a throat swab and a serum specimen from each suspected measles case to increase the sensitivity of diagnosis and enhance case classification. The sensitivity and specificity of IgM ELISA assays are influenced by the format of the test which include indirect or capture assays. False positive IgM results can result in unnecessary and costly public health interventions. False negative or negative serologic results can occur as a function of test sensitivity resulting from early (0-3 days after rash onset) specimen collection or assay interference and can cause a delay in detection or a missed opportunity to identify a true measles case. The goal of this comparison study was to identify commercially available ELISA assays that detect measles IgM with high levels of sensitivity and specificity using the CDC in-house IgM capture assay as the standard of comparison. The CDC in-house IgM capture assay was used as the comparator as it demonstrated complete concordance with molecular test results (i.e., RT-PCR) for confirmed measles cases.

## Methods

### Serum samples

A panel of 90 previously characterized and deidentified sera were chosen for use in this comparison study. To evaluate test performance against the CDC IgM capture assay, a range of detectable levels of IgM positive samples (n=32) and a group of negative samples (n=37) were selected in addition to an interference panel (n=21). The sera that were evaluated for the comparison consisted of 20 cases that were IgM-positive by the CDC IgM capture assay and confirmed by detection of measles RNA in throat swabs using RT-qPCR, 12 that were IgM-positive for measles by both the CDC IgM capture assay and by another IgM assay at either a commercial or state public health laboratory, 14 that were negative by the CDC IgM capture assay and had a negative RT-qPCR result for the throat swab, 9 that were negative by CDC IgM capture assay and IgM negative by another IgM assay at either a commercial or state public health laboratory, and 14 that were negative by the CDC IgM capture assay with discrepant positive results from another IgM assay at either a commercial or state public health laboratory. These 14 sera were from patients with a low suspicion of being a measles case and were discarded as cases. Additionally, a set of 21 commercially available plasma samples (Access Biologicals, Vista, CA) known to be potentially cross-reactive or interfering with measles IgM detection were also included. Sera were blind coded and tested by three independent operators for each ELISA assay. Sample volume for a few specimens was depleted over the course of the evaluation so not all commercial kits tested the same number of samples.

### CDC in-house IgM Capture Assay

Serum specimens were tested for measles-specific IgM using the CDC measles IgM capture EIA as described (8). Briefly, results for the CDC measles IgM ELISA were determined by comparing the optical density (O.D.) readings from the nucleoprotein positive antigen (P) wells to the O.D. of the negative control antigen (N) wells. Results were defined as “Positive” if P/N ≥ 3.0 AND P-N ≥ 0.1, “Negative” if P/N ≤ 3.0 AND P-N ≤ 0.1, or “Indeterminate” if only one of the above criteria were met.

### RT-Qpcr

Specimens with accompanying samples for virus detection were tested with a real time quantitative PCR (RT-qPCR) assay described previously (9).

### IgM Kit Descriptions

The five measles IgM ELISA assay kits that were compared included: Euroimmun Anti-Measles Virus NP IgM ELISA, (Euroimmun, Lübeck Germany catalog no. EL2610-9601-4M), Virion/Serion ELISA classic measles virus IgM (Virion/Serion, Würzburg Germany catalog no. ESR102M), Awareness Technology (AwareTech) ReQuest Measles IgM (Quest International Inc., Doral, FL catalog no. 01-190M), IBL International Measles Virus IgM micro-capture ELISA (IBL International, Hambürg Germany catalog no. RE57151), and Trinity Biotech Captia Measles IgM ELISA, (Trinity Biotech, Jamestown NY catalog no. K140455). To decrease variability, all commercial kits used in the evaluation were manufactured in the same respective commercial lots. The Trinity Biotech kit is the only measles IgM test that is FDA approved.

Euroimmun, Virion/Serion and Trinity are indirect assays whereas IBL and AwareTech are capture IgM assays. For indirect IgM ELISA tests, wells are coated directly with viral antigens, serum is applied, and antigen-specific IgM antibodies are detected using anti-IgM reporter antibodies. In contrast, IgM capture ELISA tests contain wells that are coated with anti-IgM antibody. Viral antigen is subsequently applied and detected with a specific monoclonal antibody and secondary enzyme-conjugate. Sera were tested using each kit per manufacturer’s instructions. Each commercial assay kit was also tested with the high positive, low positive and negative control plasma included as standard controls in the CDC IgM capture assay (Access Biologicals, Vista CA).

### Statistics

GraphPad Prism software (version 5.04) was used for statistical analysis and plotting of the data. Statistical significance was assigned to *p*-values <0.05 for all analyses. To validate the use of the CDC in-house capture assay as the comparator for evaluation, a Receiver Operator Characteristic (ROC) analysis comparing RT-qPCR positive and CDC IgM positive sera (n=20) to RT-qPCR negative and CDC IgM negative sera (n=23) was performed to ensure accuracy for detecting measles IgM in confirmed measles cases. After validation of the CDC in-house capture assay, ROC analysis was also used to estimate the accuracy of the commercial assays to the comparator.

## Results

### IgM Kit Comparison

Measles specific IgM was detected in all serum specimens from cases that had accompanying RT-qPCR positive results by the CDC in-house IgM capture assay in comparison to the serum specimens that had accompanying negative RT-qPCR samples. We subsequently performed ROC analysis for each commercial IgM test using the CDC IgM capture assay as the comparator based on its concordance with molecular testing results (Table 1). The average area under the ROC curve (AUC) ranged from 0.59-0.71 for all operators for each assay, indicating sufficient to moderate diagnostic accuracy with a standard error ≤0.1. The ability to accurately detect measles-specific IgM in serum specimens that had accompanying RT-qPCR-positive test results ranged from 55-83.3%, with AwareTech having the highest agreement.

**Table 1.** Performance Characteristics of Commercial IgM ELISA tests in comparison to Positive CDC IgM ELISA Samples from Cases with Positive RT-qPCR Swabs.

|  | PCR Positive/<br>CDC IgM Positive |  | ROC Analysis |  |  |
| --- | --- | --- | --- | --- | --- |
| Test System | % | Number | AUC<sup>1</sup> | Standard Error<sup>2</sup> | 95% CI<sup>3</sup> |
| CDC IgM ELISA<br>450nm (reference wavelength 600-650nm) | 100 | 20/20 | 1.00 | 0.00 | 1.000-1.000 |
| IBL International<br>450nm (reference wavelength 600-650nm) | 72.2 | 13/18 | 0.705 | 0.0824 | 0.5429-0.8691 |
| Awareness Technology, Inc.<br>450nm (reference wavelength 620-650nm) | 83.3 | 15/18 | 0.6382 | 0.0932 | 0.4555-0.8209 |
| Institut Virion/Serion<br>405nm (reference wavelength 620-690nm) | 80.0 | 16/20 | 0.5925 | 0.0916 | 0.4130-0.7720 |
| Euroimmun US, Inc.<br>450nm (reference wavelength 620-650nm) | 55.0 | 11/20 | 0.7075 | 0.0824 | 0.5459-0.8691 |
| Trinity Biotech Plc.<br>450nm (reference wavelength 600-650nm) | 55.0 | 11/20 | 0.6925 | 0.0838 | 0.5283-0.8567 |

<sup>1</sup>Area under the curve
<sup>2</sup>Standard error for AUC
<sup>3</sup>95% confidence interval

The complete panel of 69 well characterized serum specimens and serum from 21 potential interfering agents were tested by each operator with five commercial IgM ELISA assays, and results were compared to those obtained using an in-house IgM capture assay developed at the CDC (Table 2). The highest agreement with the CDC IgM ELISA occurred with the two commercial kits with capture formats (Table 2). There was an agreement of 86.2% (56/65) with IBL International, and an agreement of 85.3% (56.3/66) with AwareTech. The assay with the least agreement with the CDC in-house assay was Trinity Biotech with an agreement of 71.2% (47/66).

**Table 2.**
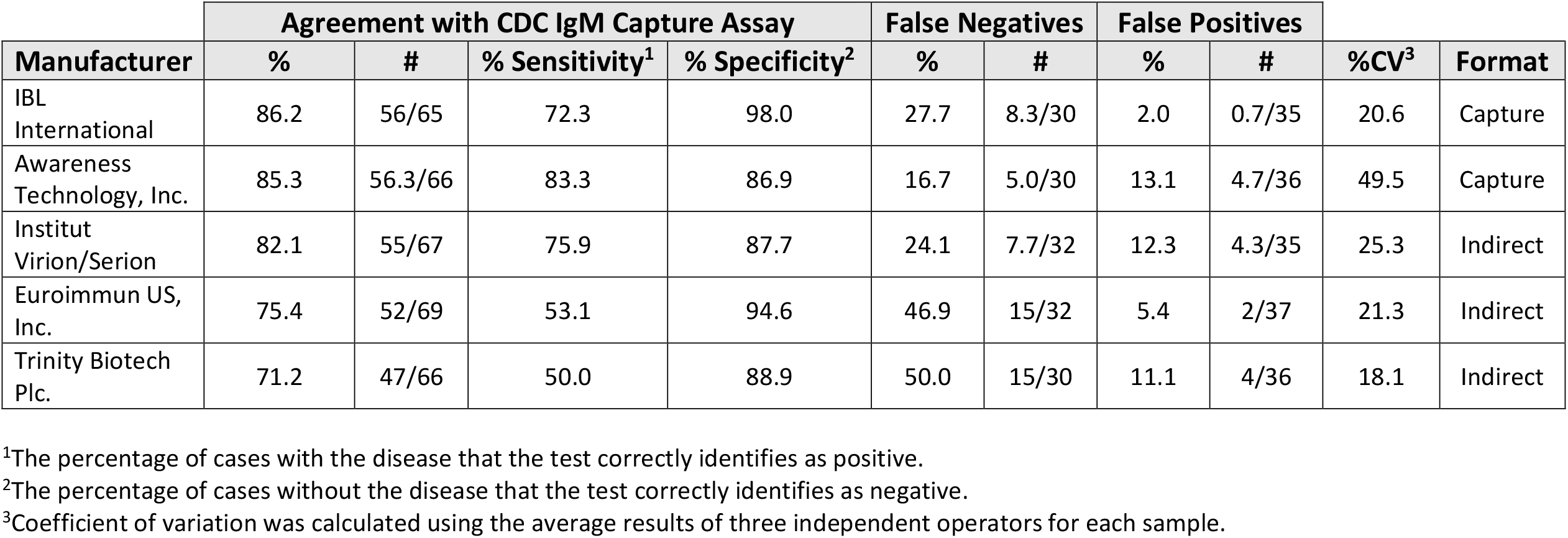
Performance Characteristics of Commercial IgM ELISA tests in comparison to the CDC IgM ELISA.

Assay sensitivity and specificity were determined using the CDC IgM assay as the comparator (Table 2). Sensitivity ranged from 50 to 83.3% for all kits tested. The commercial assay with the highest sensitivity was AwareTech with 83.3% while the lowest sensitivity at 50% occurred with the Trinity test. Specificity indicates the fraction of cases without the disease that the test correctly detects as negative. The commercial kit with the highest specificity was IBL with 98%, followed by Euroimmun with 94.6%. Specificities greater than 86% were observed in all tests. The highest number of false negatives and the failure to identify the highest number of samples that were positive by both the CDC IgM ELISA and RT-qPCR occurred in samples tested with Trinity and Euroimmun; both are indirect ELISAs. Two percent of the false positives was detected with IBL. Notably, the commercial assay with the highest coefficient of variation (CV) was AwareTech with a CV of 49.5% compared to 18.1-25.3% for the other assays.

### CDC Positive and Negative Internal Control Results

The internal controls for the CDC IgM capture ELISA (high positive, low positive, and negative control plasma) were independently evaluated on each commercial test system by all three operators. IBL, Virion/Serion, and Euroimmun assays are intended for both serum and plasma use while AwareTech and Trinity only indicate usage with serum. IBL was the only commercial assay to provide correct results with all internal controls. The in-house negative control was negative on all assays while the high control gave a positive signal only with AwareTech and Virion/Serion. Neither the high or low positive controls were positive with Euroimmun and Trinity.

### CDC IgM Test Characteristics of Samples with False Results on Commercial *Kits*

To provide a comprehensive view of kit performance, the CDC IgM capture assay test characteristics for samples with false results on commercial kits are shown in Figure 1A. Optical density values for both the positive (measles recombinant NP) and negative antigens (*Spodoptera frugiperda* cell lysate; SF9) were plotted with the cut-off criteria for the CDC IgM assay to show the relationship of the absorbance values for each antigen tested by the CDC test to the qualitative result for each commercial assay. Most of the false positive and false negative results for the IBL kit were close to the cut-offs for both positive (NP) and negative (SF9) antigens. IBL, AwareTech, and Virion/Serion had multiple false positive results with absorbance values in the lower range of detection. In contrast, Euroimmun and Trinity measured many false negatives for specimens that registered in the higher range of detection for the positive (NP) antigen on the CDC IgM ELISA. The qualitative results for all samples tested by each commercial assay are plotted in relation to the P/N and P-N values for the samples tested by the CDC IgM ELISA in relation to the cut-off criteria for the CDC IgM assay (Figure 1B). The kit with the least number of discrepant results when compared to the CDC IgM assay was IBL. The false negatives and false positives of IBL fell within the lower range of the cut-off for being positive or negative in the CDC IgM ELISA. The kits that had more false positives in comparison to the CDC IgM ELISA cut-offs were AwareTech and Virion/Serion, while Euroimmun and Trinity had higher numbers of both false negatives and false positive results compared to the CDC assay. More false positives that were clearly negative by the CDC IgM ELISA was identified with AwareTech, and Virion/Serion. Similarly, the false positives and negatives for Euroimmun and Trinity were well above and below the cut-off criteria for a negative or positive result, respectively.

**Figure 1.**
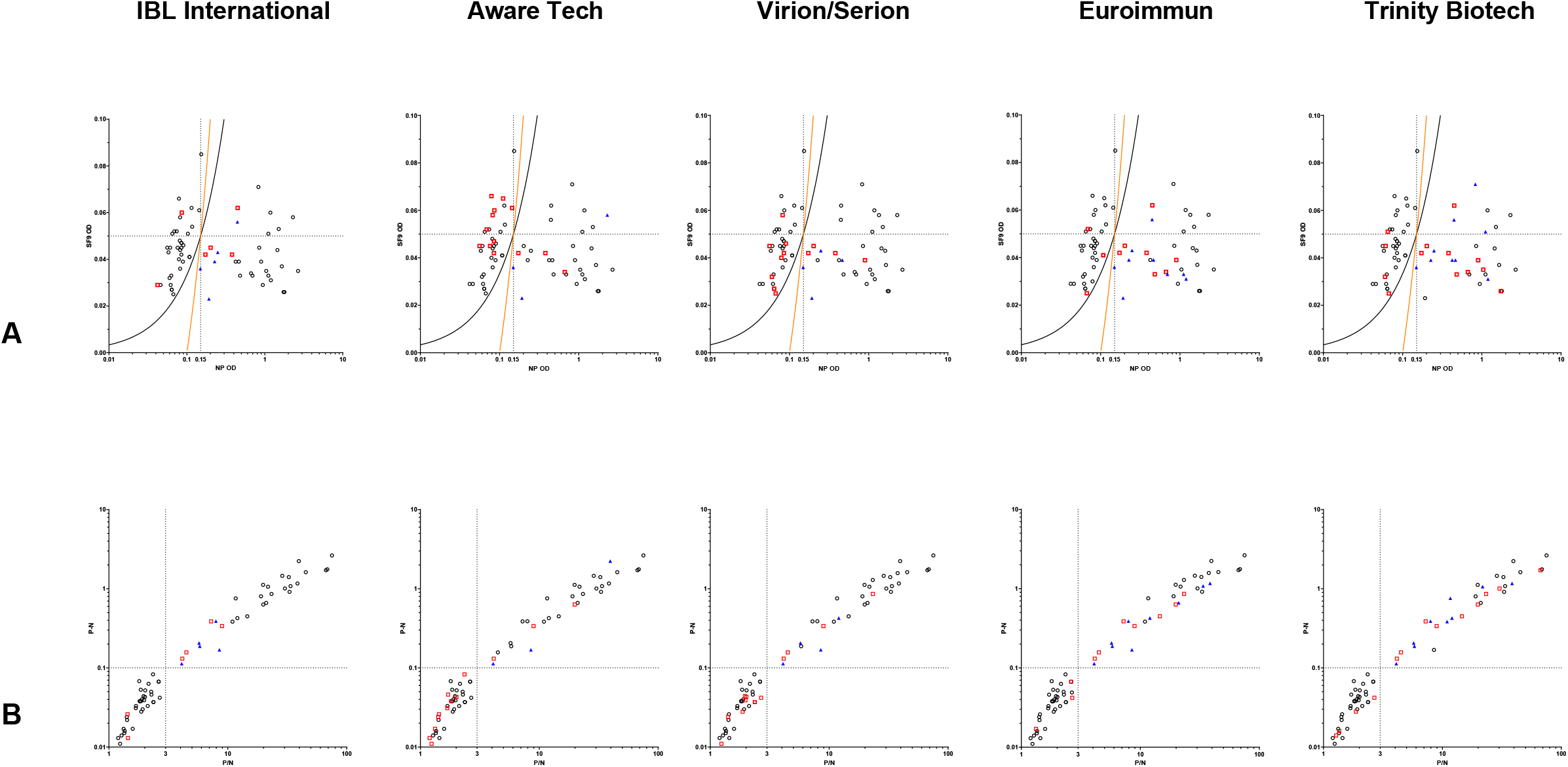
Performance of each commercial IgM assay kit relative to the CDC IgM ELISA. (A) Absorbance values for samples tested by the CDC IgM ELISA for each commercial kit. The absorbance values of the positive (measles NP antigen) and negative (SF9 control antigen) are shown for each sample used on the comparison for each commercial ELISA test. The cut-off criteria for the negative and positive results are shown by the orange line (P-N=0.1) and the black line (P/N=3.0). Black circles indicate CDC IgM results that qualitatively agree with the indicated commercial test. Red squares indicate false positive results. Blue triangles indicate false negative results that were confirmed as positive by PCR. (B) Positive and Negative cut-off values for samples tested by the CDC IgM ELISA in comparison for each commercial kit. The cut-off criteria for P/N vs. P-N values are shown by dotted lines. Black circles indicate CDC IgM results that qualitatively agree with the indicated commercial test. Red squares indicate false positive results. Blue triangles indicate false negative results that were confirmed as positive by PCR.

The cumulative false results were compared across all five test kits in relation to the number of samples tested in this evaluation. The kits with the least number of false negative results were IBL and AwareTech (14% and 15%, respectively), but out of the two AwareTech had more false positives (7%) than IBL. Trinity and Euroimmun had the highest percentage of false negatives ranging from 24.64 to 28.78% (Figure 2A). The most stringent criteria to identify measles cases are samples that have both positive CDC IgM results and positive RT-qPCR results. Euroimmun and Trinity had the highest percentage of false negatives among this group (40.6-48.3%), while the remaining 3 assays had comparable performance in identifying samples that were IgM positive and from a case with a RT-PCR positive result (Figure 2B). IBL had 5 false negatives and those samples were in the lower range of detection by the CDC IgM ELISA.

**Figure 2.**
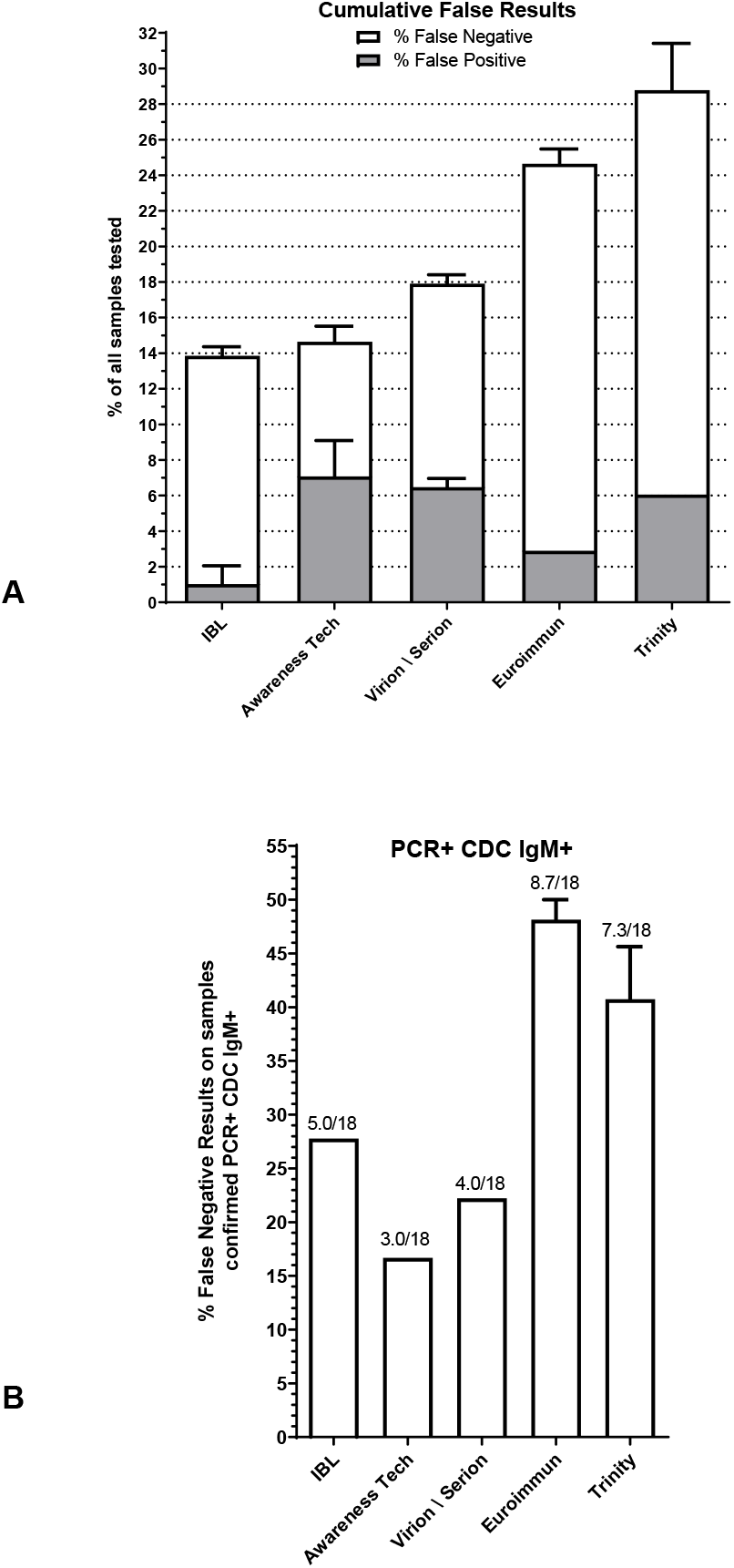
False negative and false positive IgM results from commercial ELISA kits. (A) Cumulative false negative and false positive results from each commercial ELISA were calculated based on agreement with the CDC IgM ELISA. The false results are expressed as a percentage of total samples tested. (B) The fraction of false negative results from each commercial kit are shown for cases that were IgM-positive by the CDC ELISA and confirmed by RT-PCR in accompanying throat swabs.

### Interference Panel

Twenty-one samples known to interfere with measles IgM ELISA assays were tested by three independent operators for all IgM tests. The CDC IgM ELISA is historically known to react with some specimens from individuals infected with varicella zoster (VZV) and parvovirus B-19 (10). All operators for the CDC IgM ELISA reported positive IgM results to VZV plasma and one operator measured reactivity to parvovirus B-19. Interestingly, two operators observed positive IgM results to dengue virus. IBL and Euroimmun were the only assays that did not demonstrate any interfering activity with any of the samples. There was reactivity to HSV-1/2 and parvovirus B-19 samples for all operators on Virion/Serion. Positive results to HSV-1/2 and Zika virus samples was detected by all operators, and there was operator-dependent reactivity against plasma samples positive for rheumatoid factor and Epstein-Barr virus for Trinity. The IgM results to HSV 1/2 was positive by all operators for AwareTech, and the reactivity with seven of the twenty-one samples (Parvovirus B-19, Zika virus, Lyme IgM, Rheumatoid Factor, HIV, and Enterovirus IgM) depending on the operator.

## Discussion

Detection of measles IgM in patient serum is an important component of the laboratory diagnosis of measles, and the CDC recommends collection of both a serum specimen for IgM detection and a nasopharyngeal/throat swab for RT-PCR from each suspected case to increase the likelihood of detecting measles infection. The objective of this study was to determine the performance characteristics of 5 commercially available measles IgM tests using the CDC in-house developed IgM capture assay as the comparator. Well-characterized patient sera with a wide range of positive and negative IgM values were used, and each specimen was confirmed as negative or positive by two independent test methods prior to inclusion in the test panel. Each IgM ELISA test was performed by three independent operators to provide a measure of repeatability and precision.

Clear differences in kit performance were observed amongst the commercial assays. The greatest agreement with the CDC IgM capture assay was with the two capture assays IBL and AwareTech with 86.2 and 85.3% agreement, respectively. The highest specificity at 98% (2% false positive rate) while maintaining a sensitivity of 72.3% (27.7% false negative rate) was with IBL. The variability from replicate determinations of the same sample for IBL was significantly lower (CV of 20%) than that seen for the AwareTech assay (CV of 49.5%). Higher sensitivity (83.3% vs 72.3%) was seen with AwareTech than IBL but with lower specificity (86.9% vs 98%). Of particular interest are results from the 18 specimens that were positive by the CDC IgM Capture assay and collected from cases with a positive RT-PCR result in an accompanying throat swab. Five false negatives were detected by IBL among this group compared to 3 for the AwareTech assay. The false negatives results obtained using the IBL kit were in the lower range of detection for the CDC IgM assay, as were 2 of the 3 false negatives for AwareTech. However, one of the three false negatives for AwareTech was a strong positive by the CDC IgM assay.

Of the three indirect assays, specificity was highest for Euroimmun at 94.6% compared to 87.7% for Virion/Serion and 88.9% for Trinity. The sensitivity of Euroimmun and Trinity were low at 53.1% (15/32 samples tested were false negative) and 50% (15/30 were false negative) compared to Virion/Serion which had the highest sensitivity (75.9%) of the indirect assays and the second highest sensitivity overall. When considering specimens that were IgM-positive and collected from cases with a PCR-positive throat swab, Virion/Serion had a false negative rate of 22% while Euroimmun and Trinity were almost twice that at 48.3% and 40.6%, respectively.

IBL was the only commercial assay evaluated that was able to accurately detect our in-house high positive and low positive controls. The high positive control was detectable by AwareTech and Virion/Serion but not the low positive control. Of concern, neither the high positive or low positive control was detected by the Euroimmun and Trinity assays, even though Euroimmun indicates usage with both serum and plasma. Indirect IgM assays are prone to interference from measles-specific IgG in the specimens, which could explain the inability of the Euroimmun and Trinity kits to detect positive controls.

The interference panel was not comprehensive but did include twenty-one common potentially interfering agents. As previously shown in literature, the most likely interfering agents (parvovirus B-19, dengue virus and VZV) were also reactive in the CDC IgM assay and some of the commercial assays (10). Interestingly, IBL and Euroimmun were the only assays that did not demonstrate reactivity with any of the samples. To fully examine the potential for interfering reagents, a larger number of samples for each of the interfering agents should be tested but our serum volume was limited, and we were not able to include additional specimens.

Heibert and colleagues recently evaluated the test characteristics of eight commercially available measles IgM kits, including the kits from Virion/Serion, Euroimmun, and IBL that are included in our current study (11). This report found that IgM capture assays had the best sensitivity and specificity for detecting measles IgM antibodies. IBL (an IgM capture assay) had equivalent results to the comparator, the Siemens (Engyznost) kit, which is no longer available. Heibert et al. employed a more robust cross-reactivity panel containing 187 samples, but, similar to our findings, the IBL kit was non-reactive with their cross-reactivity panel.

Evaluation and interpretation of measles IgM serologic results can be complex, particularly in measles elimination settings. In settings of low disease prevalence – such as the US – a considerable portion of positive measles test results may be false positives. Laboratories should consider the performance characteristics of the IgM test kits when developing testing algorithms to confirm measles case. Additionally, consideration of epidemiologic and clinical factors related to a case plays a critical role in interpreting laboratory results and providing confirmation of measles diagnosis.

## Data Availability

All data produced in the present study are available upon reasonable request to the authors.

## Acknowledgments

The authors would like to thank Christine Badeau and Jessica Coates for assistance with laboratory testing.

